# HIV-1 infection induces functional reprogramming of female plasmacytoid dendritic cells associated with enhanced *TLR7* expression

**DOI:** 10.1101/2022.03.14.22272210

**Authors:** Flora Abbas, Ali Youness, Pascal Azar, Claire Cenac, Pierre Delobel, Jean-Charles Guéry

## Abstract

Plasmacytoid dendritic cells (pDCs) express TLR7, a ssRNA-sensor encoded on the X chromosome, which escapes X chromosome inactivation (XCI) in females. pDCs are specialized in the production of type 1 interferons (IFN-I) through TLR7 activation which mediates both immune cell activation and also reactivation of latent HIV-1. The effect of HIV-1 infection in women under antiretroviral therapy (ART) on pDC functional responses remains poorly understood. Here, we show that pDCs from HIV/ART women exhibit exacerbated production of IFN-α and TNF-α as compared to uninfected controls (UC) upon TLR7-activation. Because *TLR7* can escape XCI in female pDCs, we measured the contribution of *TLR7* allelic expression using SNP haplotypic markers to rigorously tag the allele of origin of *TLR7* gene at single cell resolution. Herein, we provide evidence that the functional reprogramming of pDCs in HIV/ART women is associated with enhanced transcriptional activity of the *TLR7* locus from both X chromosomes, rather than differences in the frequency of *TLR7* bi-allelic cells. These data reinforce the interest in targeting the HIV-1 reservoir using TLR7 agonists in women.

## Introduction

Despite the efficacy of long-term antiretroviral therapy (ART), HIV-1 latent reservoir persists in individual living with HIV-1 and can reactivate and cause active infection upon treatment interruption (Mitchell *et al*, 2020). HIV persists in a small pool of cells, particularly memory CD4 T cells, harboring integrated and replication-competent viral genomes (Dufour *et al*, 2020). Latency removal agents (LRA) have been developed as potential means to reduce the HIV-1 reservoir by inducing reactivation of latent proviruses followed by elimination of the resulting infected cells by the immune system (Dufour *et al*., 2020). Second generations of LRAs have been developed that stimulate the endosomal Toll-like receptors TLR-7, TLR-8 or TLR-9, which are of high interest as potential cure strategies due to their capacity to both reverse HIV-1 latency and enhance HIV-1-specific immune control (Borducchi *et al*, 2016; Borducchi *et al*, 2018; Lim *et al*, 2018; Macedo *et al*, 2019; Meas *et al*, 2020). Transient reactivation of the latent SIV reservoir and sustained viral remission have been obtained using the TLR7 agonist ligand, (GS-9620), in simian immunodeficiency virus (SIV) infected rhesus macaques under ART alone (Lim *et al*., 2018) or in combination with anti-envelope antibodies or therapeutic vaccines (Borducchi *et al*., 2016; Borducchi *et al*., 2018).

Plasmacytoid dendritic cells (pDCs) express TLR7, which senses ssRNA, and induces the secretion of copious amounts of type I IFNs (IFN-I) that promote cell-autonomous antiviral defense through interferon inducible genes, and also serves as bridge to potentiate innate and adaptive immunity promoting antibody responses and enhancing cytotoxic T cells and NK cells potential (Gonzalez-Navajas *et al*, 2012). TLR7-agonist ligands can mediate both immune cell activation and HIV-1 expression in cells from HIV-1-infected individuals on suppressive ART (HIV/ART) (Tsai *et al*, 2017; Van der Sluis *et al*, 2020). In this model, the reactivation of the latent HIV-1 reservoir in PBMCs from ART-suppressed individuals was dependent on pDC-mediated production of IFN-α which was strongly induced in HIV-infected PBMCs (Tsai *et al*., 2017). Understanding the mechanisms controlling the expression of TLR7 and the production of IFN-I is therefore an important issue in HIV-1 infection, in particular for the development of new strategies for viral eradication (Borducchi *et al*., 2016; Borducchi *et al*., 2018; Lim *et al*., 2018; Macedo *et al*., 2019). Although pDC numbers are reduced in the blood compartment of HIV/ART (Fontaine *et al*, 2009; Kamga *et al*, 2005; Sabado *et al*, 2010), limited studies have been performed to assess their functional properties in response to TLR7 agonist ligands, particularly in women.

Herein, we *ex vivo* assessed the innate function of pDCs upon TLR7 stimulation by controlling for the presence of the rs179008 single nucleotide polymorphism c.32T allele which inhibits TLR7 protein expression and pDC innate functions, selectively in female, but not in male (Azar *et al*, 2020; Guery, 2021). We show here that HIV/ART women exhibited exacerbated production of IFN-α and TNF-α as compared to uninfected controls (UC). Because *TLR7* can escape X chromosome inactivation (XCI) in human female pDCs (Souyris *et al*, 2018), we then assessed the contribution of *TLR7* allelic expression on the functional reprogramming of pDCs in HIV/ART women, using SNP haplotypic markers to rigorously tag the allele of origin of *TLR7* gene at single cell resolution.

## Results and Discussion

### The *TLR7* rs179008 c.32T allele inhibits IFN-α production by pDCs from HIV-1-infected women under ART

We recently established that the *TLR7* rs179008 c.32T allele, which the frequency distribution was similar between HIV-1 infected and control women (Azar *et al*., 2020), is a sex-specific protein expression quantitative trait locus (pQTL) where c.32T allele carriage was associated with both impaired TLR7 protein expression and TLR7-driven production of IFN-α by pDCs in females, but not in males (Azar *et al*., 2020). Herein, we investigated the impact of the c.32T minor allele of rs179008 on the functional response of pDCs from HIV/ART women with sustained undetectable viral load. HIV/ART women with known A/A or A/T genotype from the ANRS EP53 X_LIBRIS cohort were reassessed for the relative frequency of circulating pDCs and the functional response of their blood pDCs following stimulation with TLR7 ligands using the flow cytometry strategy described in Fig EV1 as previously described (Azar *et al*., 2020). We observed a lower frequency of pDCs in the blood of HIV/ART women relative to age-matched uninfected controls (UCs) (Fig 1A), as per previous reports (Fontaine *et al*., 2009; Kamga *et al*., 2005; Sabado *et al*., 2010). Among HIV/ART and UC women, pDC frequencies in the AA and AT subjects were equivalent (Fig 1B). Fresh PBMCs were stimulated with DOTAP vesicles loaded with Gag_RNA1166,_ a synthetic HIV-1-derived RNA ligand of TLR7 and TLR8. In agreement with our previous work with healthy female donors, the frequency of IFN-α-producing pDCs in Gag_RNA1166_-stimulated PBMCs was significantly reduced in AT heterozygous patients compared to AA HIV/ART women (Fig 1C). Thus, the rs179008 c.32A>T pQTL is also a functional polymorphism controlling TLR7-driven production of IFN-I by pDCs in HIV-1/ART women.

**Figure 1:**
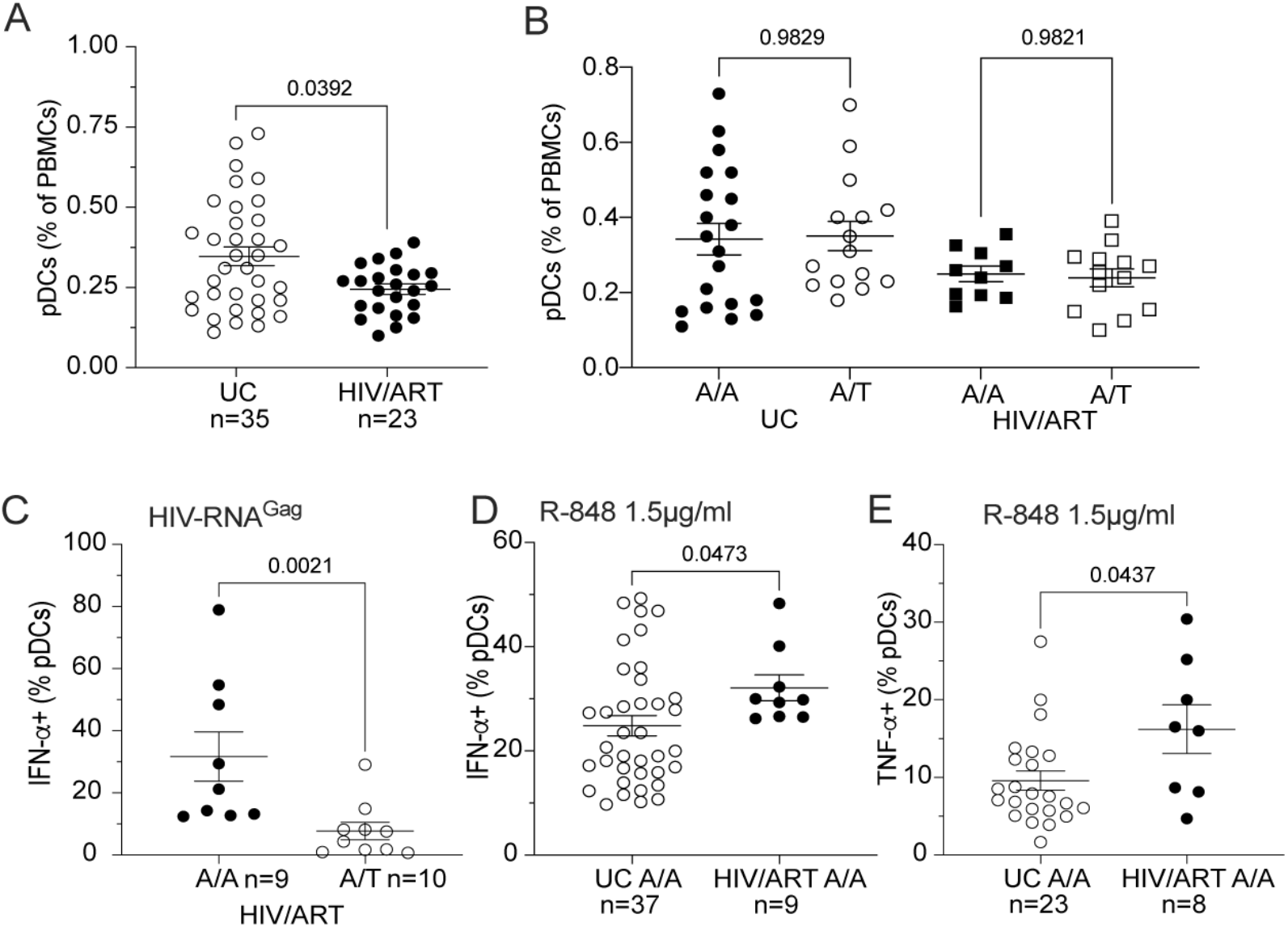
pDCs from HIV/ART women exhibited increased functional responses to TLR7 ligands compared to UCs. A PBMCs from HIV/ART females and age-matched uninfected controls (UCs) were monitored for the percentage of pDCs (lin^-^ CD123^+^BDCA4^+^) as indicated in Supporting information Fig EV1. B The frequency of blood pDCs are represented according to the expression of the indicated genotype of the rs179008 SNP (A/A vs A/T) of *TLR7*. C Fresh PBMCs from the HIV-1^+^ women with the indicated rs179008 genotypes were stimulated with HIV-derived Gag_RNA1166_ in DOTAP and intracellularly stained for IFN-α expression in CD123^+^BDCA4^+^ pDCs. D, E Fresh PBMCs from HIV-1/ART and UC females homozygous for the *TLR7* rs179008 A/A SNP were stimulated with TLR7/8 ligand R-848 (1.5 µg/ml), and intracellularly stained for IFN-α and TNF-α. Frequencies of pDCs producing IFN-α (D) or TNF-α (E) are shown. Data information: (A-E) Errors bars represent the mean ± SEM. Data from individual subjects are shown. Statistical analysis was performed using the Mann-Whitney test. *P* values are shown.

### Chronic HIV-1 infection is associated with enhanced functional state of TLR7 stimulated pDCs in HIV/ART women carrying the rs179008 AA SNPs

As TLR7-driven functional response of pDCs has been reported to remain intact under ART (Chang *et al*, 2012, Tsai, 2017 #4699), we next compared the frequency of IFN-α and TNF-α producing pDCs in response to optimal stimulation with the TLR7/8 ligand Resiquimod (R-848) in HIV/ART women compared to age-matched UCs, both homozygous for the frequent rs179008 A allele. As shown in Fig 1D and E, pDCs from HIV/ART females or UCs were efficiently stimulated to produce IFN-α and TNF-α. Of note the average frequency of IFN-α^+^ pDCs was significantly higher in HIV/ART females compared to uninfected controls (Fig 1D). The same trend was found for the TNF-α response (Fig 1E) suggesting that all pathways downstream of TLR7-signaling were substantially increased in HIV/ART women.

### X chromosome inactivation escape of TLR7 in pDCs is similar between HIV/ART and UC women

*TLR7* is encoded on the X chromosome and has been shown to escape XCI in a substantial proportion of immune cells from healthy women (Hagen *et al*, 2020; Souyris *et al*., 2018). However, whether the status of XCI escape can be affected by chronic HIV-1 infection has never been examined. Because the escape from XCI of the *TLR7* gene has been found associated with enhanced functional characteristics in woman B cells (Souyris *et al*., 2018) and pDCs (Hagen *et al*., 2020), we decided to design a single-cell RT-qPCR approach to measure the frequency of *TLR7* biallelic cells among pDCs from HIV/ART women. We tested the hypothesis that the enhanced TLR7-driven responsiveness of pDCs in this population could be associated with a higher frequency of cells with escape from XCI of *TLR7* gene compared to uninfected subjects. The experimental work flow is depicted in Fig EV2. To tag the expression of *TLR7* gene, we used heterozygous female donors expressing the rs179008 A/T and the rs3853839 G/C SNPs (Fig 2A, Fig EV2) as previously reported (Souyris *et al*., 2018). Frozen PBMCs were cultured overnight in the presence of IFN-ß as we found that this was associated with enhanced pDC recovery compared to the condition without cytokine (Fig EV3). Moreover, type-1 IFN signaling has been suggested to promote optimal expression of TLR7 at single cell resolution (Hagen *et al*., 2020). pDCs were single-cell sorted into two 96 well-plates in order to have at least between 100-200 pDCs analyzed in this assay using the gating strategy shown in Fig EV3. Indeed, we found that the cumulative count of single-cell sorted pDCs required to achieve robust determination of allelic expression of *TLR7* was already achieved at cell count ≥ 100 cells (Fig EV2C and D). By using the ratio of allele-specific fluorescence signals in the KASP PCR (Fig EV2B), we were able to measure the relative abundance of *TLR7* transcripts derived from either allele on each chromosome at a single cell level. Each cell was then classified into mono- or bi-allelic cell for *TLR7* expression when the minor allele frequency was estimated to range below or above 10 % of the relative proportion of *TLR7* transcripts, respectively (Fig EV2B). The allelic expression of *TLR7* using the simultaneous determination of both SNPs from each single-cell from two double-heterozygous female donors bearing distinct haplotypes are shown in Fig 2B-D. The allele-of-origin profiles were plotted according to the expression of the rs3853839 3’UTR SNP into mono-allelic C or G cells and bi-allelic GC cells (Fig 2C-E). Within each subset, we then reported the results regarding allelic expression of the second rs179008 A/T SNPs to determine the haplotypic association of both SNPs. For the female subject bearing the A-C/T-G haplotype (Fig 2 B and C), the majority of cells with mono-allelic expression of C or G allele of rs3853839 were also positive for either the A or the T allele of rs179008, respectively, indicating that the A-C and the T-G SNPs were located on the same X chromosome (Fig 2C). However, some single cells (<5%) were also positive for the SNPs belonging to the alternative X chromosome and were reassigned as bi-allelic cells. Similarly for the second female subject, using the same strategy we established that this female donor was bearing the T-C/A-G haplotype (Fig 2 D and E). Haplotype inference and allelic reassignment resulting in the final determination of haplotype profiling for all donors are shown in Fig 2 F and G. Using this approach, we found a significant underestimation of bi-allelic cell frequency when each single SNP were independently used for allele of origin measurement compared to the joined analysis of the diplotype SNPs (Fig 2G). By contrast, no significant differences in mono-allelic cell frequencies were observed using each SNP individually or as haplotype (Fig. 2F). Together, these results demonstrate that the parallel analyses of two SNPs from the same double-heterozygous female donors give an accurate and concordant estimation of the frequency of cells with mono- and bi-allelic expression of *TLR7*.

**Figure 2:**
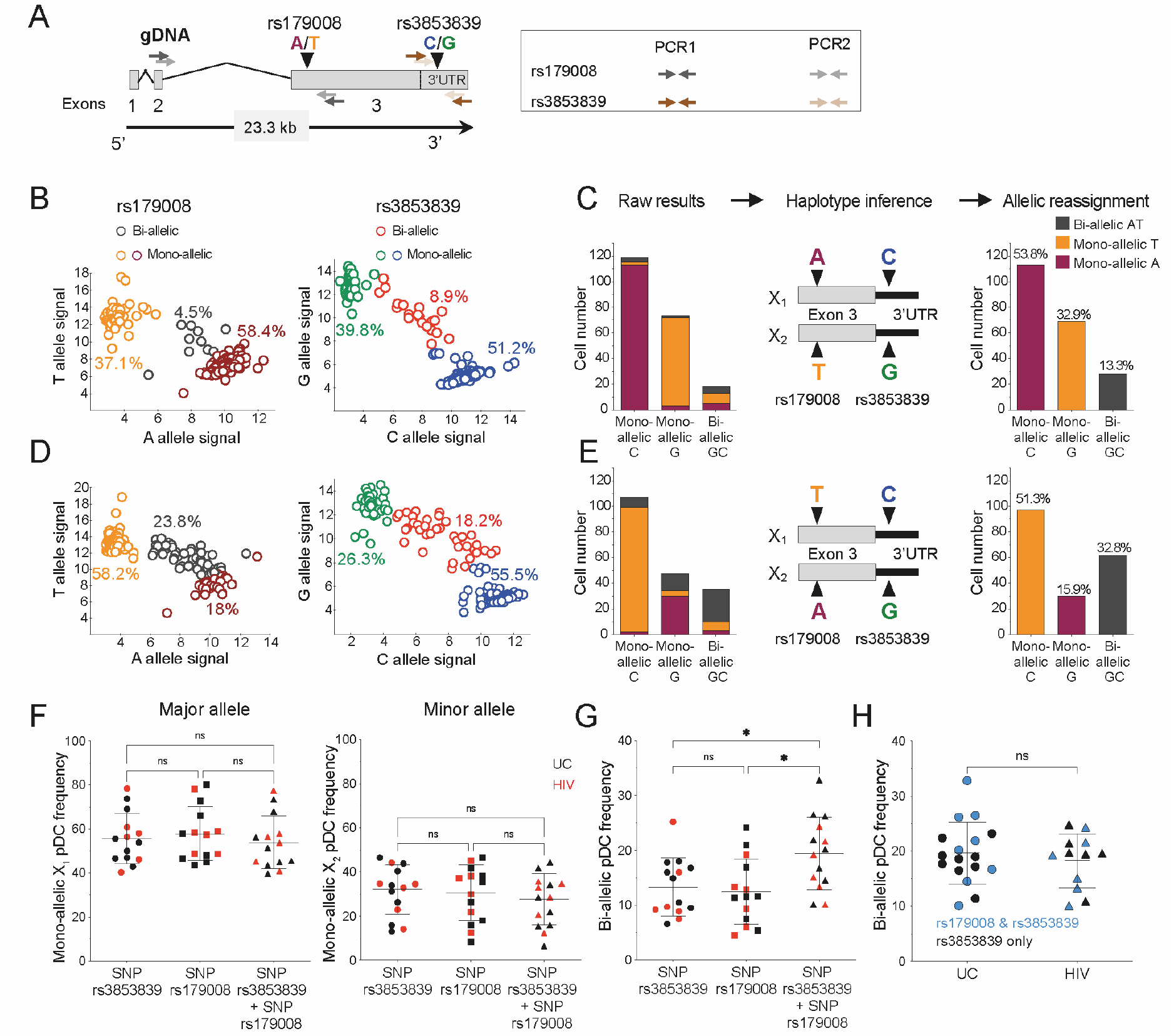
The frequencies of pDCs with biallelic expression of *TLR7* are similar between HIV/ART and UC women. A Map of *TLR7* locus with SNPs rs179008 (A/T) and rs3853839 (C/G) and the positioning of the primers used for sequential PCR amplification of *TLR7* cDNA at single-cell resolution. B, D Representative allele-of-origin profile of individual pDCs for the SNPs *TLR7* rs3853839 (right panel) and rs179008 (left panel) from 2 different heterozygous female donors. C, E Proportion of pDCs monoallelic A, T or biallelic AT pDCs within monoallelic C, G or biallelic GC for *TLR7* from two representative heterozygous women, bearing either the A-C/T-G (B, C) or the T-C/A-G (D, E) haplotypes. Raw results, haplotype inference and cell reassignment are shown in (C, E). F, G Frequencies of mono-allelic (F) or biallelic (G) pDCs from healthy (n=8) and HIV-1^+^ (n=6) women by using the SNP rs3853839 and SNP rs179008 alone or in combination. H Comparison of *TLR7* bi-allelic cell frequencies in pDCs from UCs and HIV/ART individual subjects determined using the SNP haplotypes or rs3853839 G/C SNP only. Data information: (B-E) pDCs were single-cell sorted from two representative women heterozygous for both SNPs, displaying various proportion of biallelic cells; each dot represents one cell with monoallelic or biallelic expression of *TLR7*. (F-H) Errors bars represent the mean ± SEM. (F, G) Statistical analysis was performed using a one-way ANOVA followed by a Tukey’s multiple comparisons test. ns, not significant. Exact P values are shown. (H) Frequencies of *TLR7* bi-allelic pDCs from UCs or HIV/ART determined using the TLR7 SNPs rs3853839 and rs179008 haplotypes, or the rs3853839 SNP G/C only. Mann-Whitney test was used for statistical analysis. ns, not significant.

We then compared the frequency of cells with bi-allelic expression of *TLR7* in single-cell sorted pDCs from female heterozygous for the rs179008 and rs3853838 SNPs (UC n=8; HIV/ART n=6). The frequencies of pDCs with bi-allelic expression of *TLR7* were ranging from 8 to 32% in the 14 females tested, and no significant differences were observed between UC and HIV/ART women (Fig 2G and H). We also analyzed the frequency of bi-allelic cells in females heterozygous for the rs3853839 G/C SNP. Again, using this single SNP to tag allelic expression, we also found similar frequencies of bi-allelic cells in pDCs from HIV-1 infected women compared to UC women (Fig 2H). Combined, these results represent the largest quantitative assessment of the frequency of *TLR7* biallelic cells in human immune cells with 29 subjects examined in total. In agreement with previous works (Hagen *et al*., 2020; Souyris *et al*., 2018), this demonstrates that escape from XCI of *TLR7* is a common and highly reproducible feature in female pDCs not only in UC but also in HIV/ART women. Differences in the frequency of pDCs with biallelic expression of TLR7 are unlikely to explain the enhanced functional responses observed in HIV/ART women (Fig 1).

### Chronic HIV-1 infection is associated with higher expression levels of TLR7 mRNA at single cell resolution

Because our experimental workflow allows us to quantify the mRNA expression level from each allele of *TLR7* (Fig EV2A), we compared the expression levels of *TLR7* mRNA expressed from each X chromosome in mono- and bi-allelic cells in single-cell sorted pDCs. In mono-allelic cells, *TLR7* mRNA relative expressions were similar between the two X chromosomes in all the donors, regardless of the expression of the 179008 and rs3853839 SNPs. This was observed not only in UCs, but also in HIV/ART women, whatever the haplotype born (Fig EV4A and B). These results demonstrate, at single-cell resolution, that carriage of one or the other of the minor allele rs179008 A/T or the rs3853839 G/C had no measurable impact on *TLR7* mRNA expression in female pDCs (Fig EV4C and D). This was expected for rs179008 T allele from our previous work (Azar *et al*., 2020), but much less for rs3853839 C allele which was previously reported to control the binding of the microRNA-3148 and *TLR7 mRNA* degradation (Deng *et al*, 2013). Our results now show that miR-3148 binding to *TLR7* 3’-UTR segment bearing the C allele is unlikely to control *TLR7* mRNA level in female pDCs. Our results also indicate that the expression of *TLR7* from the active X chromosome (Xa) is similar in the mosaic population of mono-allelic pDCs whatever the parent-of-origin of the Xa (Xm or Xp), contrary to what has been suggested in mice (Golden *et al*, 2019). We therefore decided to pool the data from the mono-allelic populations and compared them to the *TLR7* expression level detected in the TLR7 biallelic cells. Both in UCs and in HIV/ART women, we found significantly higher *TLR7* mRNA transcript levels in pDCs with biallelic than those with mono-allelic *TLR7* expression (Fig 3). Although, these differences were low (1.13- to 1.14-fold in UCs; 1.25 to 1.33-fold in HIV/ART), they were consistent with expression of this gene from both X chromosomes, including the Xi, in agreement with previous works in B cells (Souyris *et al*., 2018) and pDCs (Hagen *et al*., 2020).

**Figure 3:**
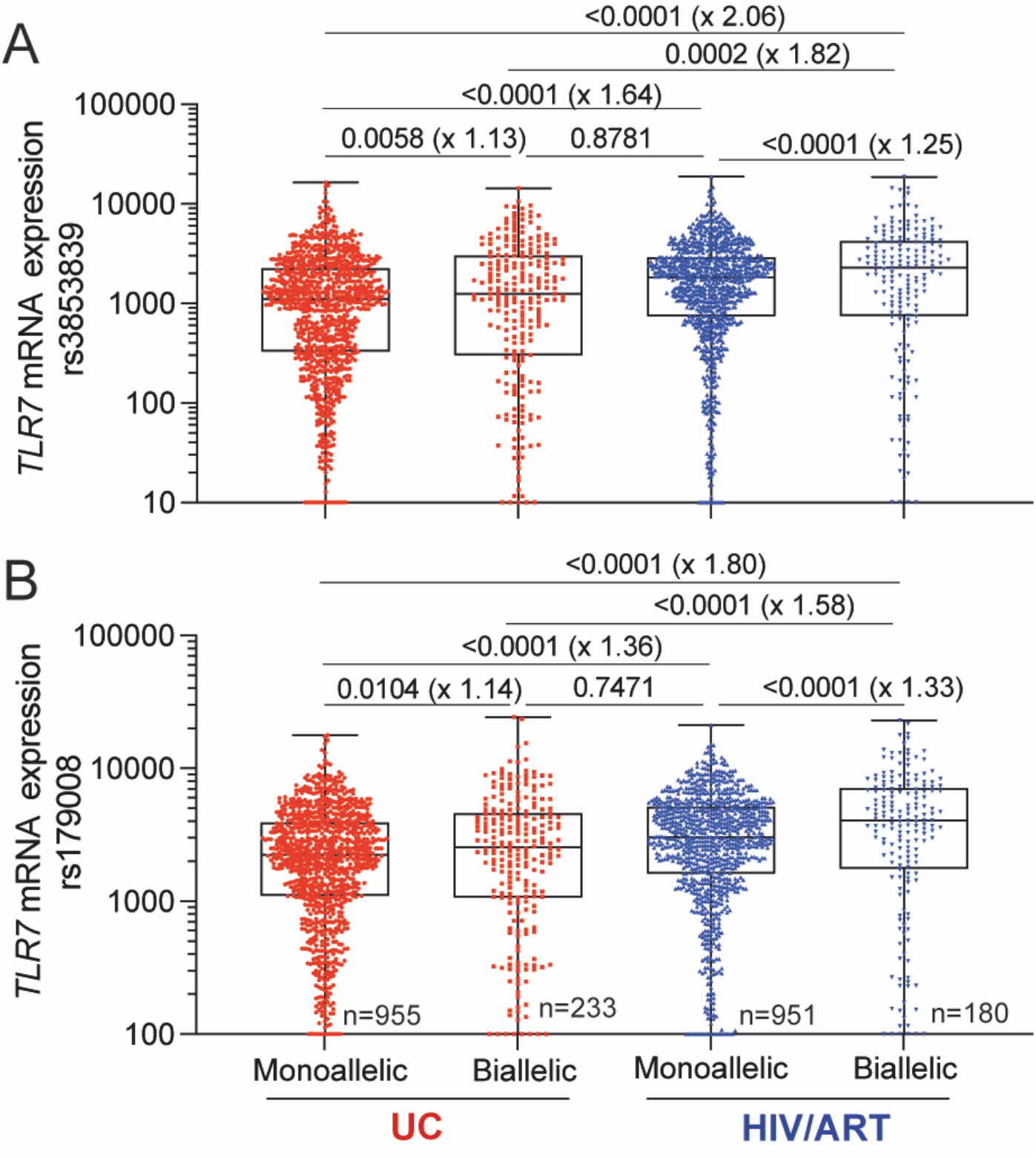
Single-cell analysis reveals enhanced *TLR7* mRNA expression in pDCs from HIV-infected women under ART. A, B Relative expression of *TLR7* mRNA in pDCs from UCs or HIV/ART women with either mono-allelic or bi-allelic expression of *TLR7* were calculated at single-cell resolution using primers specific for either the rs3893839 (A) or the rs179008 (B) cDNA sequence of *TLR7* as shown in Fig EV2 and Fig 2. Data information: A, B) *TLR7* mRNA relative expression from pooled monoallelic or biallelic pDCs from UCs (n=8, n = 1188, mean cell number/donor = 146) or HIV/ART (n=6, n=1131; mean cell number/donor = 189) females. Statistical analysis was performed using a One-way ANOVA followed by a Tukey’s post-test corrected for multiple comparison. *P* values and fold-change in median values in parentheses are shown.

Unexpectedly, we observed that the relative expression of *TLR7* at single-cell resolution was substantially higher in pDCs from HIV/ART females compared to UCs regardless of the SNP studied (Fig 3A and B). The average fold-change was calculated and we estimated that *TLR7* gene expression from the Xa (mono-allelic cells) was 1.5-fold higher in average in pDCs from HIV/ART women. A similar trend was also observed in bi-allelic pDCs with higher expression levels of *TLR7* transcripts (1.58- to 1.8-fold higher) in bi-allelic cells in HIV/ART females (Fig 3 A and B). Of note, the relative expression of *TLR7* mRNA in mono-allelic pDCs from HIV/ART females was similar to bi-allelic pDCs from HCs. Together, these results provide evidence that the functional reprogramming of TLR7-driven pDC responsiveness is associated with enhanced transcriptional activity of the *TLR7* locus from the Xa, in HIV/ART women. Moreover, by comparing *TLR7* bi-allelic cells togethers, the fold-change was even higher in HIV/ART pDCs compared to the mono-allelic cell comparison. This difference was calculated and represented 1.18- to 1.2-fold increase attributable to the additional *TLR7* mRNA expression from the Xi.

Together, our results show that although pDC number is lower in HIV/ART women, frequencies of IFN-α or TNF-α producing pDCs after TLR7 stimulation are higher than in healthy women. The analysis of *TLR7* XCI escape by single cell RT-qPCR with SNPs markers, demonstrated that the frequency of *TLR7* bi-allelic pDCs were similar between HIV/ART and UC women. *TLR7* bi-allelic pDCs expressed 1.13 to 1.25-time more *TLR7* mRNA than mono-allelic pDCs. This enhanced expression of *TLR7* mRNA transcripts in biallelic pDCs compared to mono-allelic ones was more pronounced in HIV/ART females than in HCs. Surprisingly, consistent with the increased innate cytokine responses of pDC from HIV/ART females, we observed a significant upregulation of *TLR7* gene expression in pDCs including mono-allelic ones where *TLR7* was expressed almost exclusively from the Xa. These results provide evidence for a functional reprogramming of TLR7-driven pDC responsiveness associated with enhanced transcriptional activity of the *TLR7* locus on both the Xa, and also possibly from the Xi, in HIV/ART females.

It has been recently reported at single-cell resolution that female pDCs with escape from XCI in *TLR7* expressed not only higher levels of *TLR7* mRNA but also higher levels of transcripts coding for all IFN-α subtypes and IFN-ß at steady state (Hagen *et al*., 2020). This observation suggested that the expression of basal level of IFN-I mRNA could be a consequence of a higher expression *TLR7* mRNA thereby discriminating functionally mono-allelic and biallelic pDCs (Hagen *et al*., 2020). Indeed, constitutive production of low levels of IFN-I in pDCs have been reported to act in an autocrine/paracrine manner to drive high levels production of IFN-I by pDCs (Kim *et al*, 2014). Thus, pDCs with high level expression of *TLR7* mRNA due to the cell-autonomous action of XCI escape of *TLR7* (Hagen *et al*., 2020; Souyris *et al*., 2018), could belong to a group of early responder pDCs with superior ability to produce IFN-I (Wimmers *et al*, 2018). This mechanism together with the enhanced transcriptional activity of the *TLR7* locus from the Xa observed in the general monoallelic population in HIV/ART women could contribute to the functional reprogramming of pDCs in these subjects, by increasing the frequencies of pDCs with enhanced propensity to functionally respond to TLR7 agonist ligands. Of note, we found that the expression levels of *TLR7* mRNA in mono-allelic pDCs from HIV/ART females was similar to the levels measured in *TLR7* bi-allelic cells from UCs.

Our data corroborate recent observations where increased production of pro-inflammatory cytokines has been reported in monocytes stimulated through TLR4 or TLR7 in a large cohort of HIV-1 infected subjects under ART (van der Heijden *et al*, 2021). In this study the authors observed a markedly increased monocytes-derived cytokine response, mainly affecting IL-1ß, upon stimulation with LPS, imiquimod and *Mycobaterium tuberculosis* (van der Heijden *et al*., 2021). It was suggested that immune dysfunction and persistent inflammation in HIV/ART subjects due to microbial translocation or continuing HIV replication could promote functional adaptation of innate immune cells by epigenetic or metabolic reprogramming, a process known as “trained immunity” (Netea *et al*, 2020). Although epigenetic modifications have not been documented in HIV-infected subjects, future studies are warranted to investigate the mechanisms underlying the trained immunity phenotype reported in monocytes (van der Heijden *et al*., 2021) and in pDCs (our present work).

The limitation of our study is that *TLR7* expression and pDC innate function was investigated in one sex. Whether our conclusion can be extrapolated to males will deserve dedicated studies. Of note, in the study by van der Heijden et al., the majority of HIV-1-infected subjects were males, and transcriptomic profiling of monocytes revealed broad upregulation of inflammatory pathways, including enhanced *TLR7* mRNA (van der Heijden *et al*., 2021). Thus, the trained-immunity phenotype observed in male monocytes in this study could also possibly apply to male pDCs. However, it is important to note that sex differences exist regarding TLR7 protein expression and type I IFN production by pDCs, with reduced expression of both parameters in male cells compared to females (Azar *et al*., 2020; Meier *et al*, 2009; Seillet *et al*, 2012; Souyris *et al*., 2018). Whether this sex-bias in TLR7-driven pDC responsiveness could alter the trained-immunity like phenotype we currently observed is still an open question. However, because the enhanced *TLR7* mRNA expression in pDCs from HIV/ART females was not affected by the mono-allelic expression of T allele of rs179008 T SNP we can conclude that the functional expression of TLR7 protein is unlikely to contribute to the enhanced transcriptional activity of the *TLR7* locus and the trained immunity-like phenotype we observed. We believe that epigenetic modifications at the *TLR7* locus in pDCs could be induced by pro-inflammatory cytokines, including IFN-I, which could be induced at low levels upon HIV-1 provirus reactivation whatever the sex (Kamada *et al*, 2018; Mitchell *et al*., 2020). Investigating the mechanisms underlying the functional reprogramming of pDCs will be warranted but highly challenging due to the scarcity of pDCs particularly in HIV-infected subjects. Besides myeloid cells, trained immunity have been reported in lymphoid cells, including NK cells, ILC1, ILC2 and DCs reviewed in (Netea *et al*., 2020). Our study is the first to report functional reprogramming of human pDCs and could have important implication for the development of strategies targeting the HIV-1 latent reservoir by using TLR7 agonists in HIV/ART women.

## Concluding remarks

In sum, we propose that in HIV/ART women, the innate function of pDCs is increased, and is associated with markedly enhanced expression of the *TLR7* locus from the both X chromosomes. Based on previous works by others showing strong association between *TLR7* mRNA expression and enhanced propensity of female pDCs to transcribe all IFN-I family members (Hagen *et al*., 2020), we believe that enhanced transcriptional regulation of *TLR7* mRNA could contribute to this trained immunity-like phenotype through mechanisms that will deserve further investigation. Our data strengthen the interest of targeting the HIV-1 latent reservoir by using TLR7 agonists in HIV-1-infected women under ART, and suggest that it will be critical to control for the expression of the rs179008 as this SNP could be associated with blunted pDC innate functions that still persist in HIV/ART women.

## Materials and Methods

### Donors and ethical compliance

Our study is in agreement with applicable French regulations and with the ethical principles of the Declaration of Helsinki. Peripheral blood mononuclear cells (PBMCs) of healthy blood from anonymous donors at the Toulouse blood transfusion center (Etablissement Français du Sang) were from a biobank authorized under agreement number 2-15-36 by the competent ethics board, Comité de Protection des Personnes Sud-Ouest et Outre-Mer II, Toulouse. The cohort ANRS EP_53 X-LIBRIS (coordinator: Pr. P. Delobel, CHU Purpan) allows the study of HIV-1 donors has been previously described (Azar *et al*., 2020). The X-LIBRIS cohort was previously registered with ClinicalTrials.gov under identifier NCT01952587.

### Cell culture

Frozen PBMC were defrosted and washed twice in R10 media which is: complete RPMI 1640 medium supplemented with 2 mM L-Glutamine, 1 mM sodium pyruvate, 100 U/ml penicillin/streptomycin, non-essential amino-acids, 50 μM 2-mercaptoethanol (all from Invitrogen) to which 10% heat inactivated foetal bovine serum (FBS) (Sigma) was added. Cells were cultured overnight 37°C in 5% CO_2_ air incubator. For single-cell RT-qPCR KASP, after thawing, cells were cultured overnight with IFN-β 1ng/ml in R10 complete medium.

### Cell stimulation analysis of cytokines production

Fresh PBMCs (2.5 × 10^6^ cells) were seeded in 24-well plate and stimulated either with 30µg/ml HIV-1-derived Gag_RNA1166_ synthetic oligoribonucletides (Eurogentec) complexed with DOTAP (Roche) or 1.5µg/ml R-848 (Invivogen) for five hours. Brefeldin A (eBiosciences) was added for the three last hours of culture. PBMCs were surface labeled with anti-Lin-FITC (lin1, BD Biosciences), anti-BDCA4-APC (clone REA774, Miltenyi Biotec) and anti-CD123-PECy5 (clone 9R5, BD Biosciences). Cells were fixed in 2% paraformaldehyde and permeabilized in 0,5% saponin. Intracellular staining was performed using anti-IFNα-PE (clone LT27:295, Miltenyi biotech) and anti-TNFα-AF700 (clone MAb11, BD Biosciences). Flow cytometry analysis were performed on LSRII instrument (BD Biosciences). Data were analyzed using the FlowJo software V10 (Tree Star).

### Flow cytometry cell sorting

Cells were incubated with Anti-human CD32 (Fc gamma RII) (Stemcell) for 5 min at 4°C. For cell surface markers, cells were stained in MACS buffer (PBS with 1% FBS, 2mM EDTA) for 30 min at 4°C with the following antibodies: CD14-VioBlue (clone REA599, Miltenyi Biotec), CD19-PeVio615 (clone LT19, Miltenyi Biotec), BDCA4-APC (clone REA774, Miltenyi Biotec), CD123-PeVio770 (clone AC145, Miltenyi Biotec). Then cells were stained for viability with DRAQ7™ (abcam) for 5 min at 4°C. pDCs were sorted with a FACSAria II or FACS Aria-Fusion cell sorter (BD Biosciences) at one cell by well. Data were analyzed with the FlowJo software V10 (Tree Star).

### Genotyping and single-cell analysis of *TLR7* allelic expression

The workflow for single-cell cDNA analysis previously described in (Souyris *et al*., 2018) has been optimized further and is summarized in Fig EV2. Briefly, pDCs from women heterozygous for the *TLR7* SNP rs3853839 and rs179008 were single cell sorted with a FACS Aria-Fusion cell sorter (BD Biosciences) in a 96-well plate preloaded with a medium containing 2% Triton X-100, 1 U/µl RNaseOut recombinant ribonuclease inhibitor (Thermo Fisher Scientific), 940 µM dNTPs, and 12.5 ng/µl random hexamer primers (Thermo Fisher Scientific). After cell sorting, single cell lysates were subjected to RNA reverse transcription using 6.25 U/well Maxima H Minus reverse transcriptase (Thermo Fisher Scientific). Prior to KASP genotyping, both target SNPs were PCR-amplified using *TLR7* cDNA-specific primer pairs (Table S1), and negative wells screened out by real-time quantitative PCR (RT-qPCR) with nested primers (Table EV1) and SsoAdvanced Universal SYBR Green Supermix (Bio-Rad Laboratories). The qPCR and KASP fluorescence-based assays were performed using the automated pipetting system epMotion® 5070 (Eppendorf), and a Light Cycler 480 instrument (Roche). Relative allelic expression was calculated from the ratio of the FAM (allele C or A) and HEX (allele G or T) fluorescence signals from the respective KASP probes; a 4-parameter standard curve was generated regularly for each Light Cycler 480 unit, using an R script based on package *drc* as described (Souyris *et al*., 2018). Bi-allelic *TLR7* expression in a cell was inferred when the relative expression values of *TLR7* transcripts bearing the rs3853839 G allele or rs179008 T allele was comprised between 10% and 90%. A limit of detection (LOD) was defined as a Ct value of 23, then, all Ct values higher than the LOD were removed from the analysis. mRNA expression levels were defined as 2^(LOD-Ct)^ as described (Hagen *et al*., 2020)

### Statistical analysis

Statistical analyses and data presentation were performed using GraphPad Prism 9 software (La Jolla). Statistically significant differences between 2 groups were determined using two-tailed Mann-Whitney when indicated. Multiple comparisons were performed using one-way ANOVA test followed by a Tukey’s post-test corrected for multiple comparisons as indicated. Values are reported as individual values, and plotted as mean±SEM, or as median and IQ range. All statistical tests were two-tailed and p-values <0.05 were considered to be statistically significant.

## Supporting information

Supporting Informations

## Data Availability

All data produced in the present work are contained in the manuscript

## List of abbreviations

HIV-1: Human Immunodeficiency Virus-1
HIV-1/ART: HIV-1-infected female under antiretroviral therapy
TLR7: Toll-like receptor 7
pDCs: plasmacytoid dendritic cells
UC: uninfected controls
XCI: X chromosome inactivation
Xa: active X chromosome
Xi: inactive X chromosome

## Acknowledgments

We gratefully acknowledge support from F. L’Faqihi, A.L. Iscache, V. Duplan, Lidia De La Fuente, and Hugo Garnier at the flow cytometry facility (INSERM U1291, INFINITY); P. E. Paulet at the Immunomonitoring core facility (INSERM U1291, INFINITY). The technical assistance of M. Requena, M. Cazabat and R. Carcénac (Toulouse University Hospital, INSERM U1291, INFINITY) is also greatly acknowledged. This work was supported by grants from the French National Agency for Research on AIDS and Viral Hepatitis (ANRS, EP-53 study), and SIDACTION (Grant 2018-1-AEQ-12035). PA and AY were supported by fellowships from SIDACTION. AY was also supported by PhD fellowships from the CSL Behring Research Funds, Fondation des Treilles and from the “Association de la Charité des jeunes de Kafarsir (Liban).

## Author contributions

PD and JCG conceived and designed the study. FA, AY, PA, CC performed experiments. FA, AY and PA designed the single cell RT-PCR-KASP assay. FA, AY, CC and JCG analyzed and interpreted the data. PD recruited HCs and HIV/ART women within the ANRS ER_53 X_Libris cohort. FA, CC and JCG prepared the manuscript with input from their co-authors. All authors read and approved the final version of the manuscript.

