## Supporting Informations for "HIV-1 infection induces functional reprogramming of female plasmacytoid dendritic cells associated with enhanced *TLR7* expression"

### Expended View Table and Figures

**Table EV1: PCR Primer pairs used for PCR1 and PCR2 in Fig EV2A and in Fig 2A**

| Primer pair | Sequences 5'-3' | Amplimer size |
|---|---|---|
| rs3853839, pre-KASP | ACTCAGTCAGCTTCTTAAC<br>GGATACAGTACTTTGCAGT | 303 pb |
| rs3853839, real-time PCR | TCAGTCAGCTTCTTAACCA<br>CTATTTGTAGGTGGACCAT | 200 pb |
| rs179008, pre-KASP | CTTGGCACCTCTCATGCTCT<br>CTGTGCAGTCCACGATCACA | 225 pb |
| rs179008, real-time PCR | CTGCTCTCTTCAACCAGACCT<br>AAACCATCTAGCCCCAAGGAG | 140 pb |

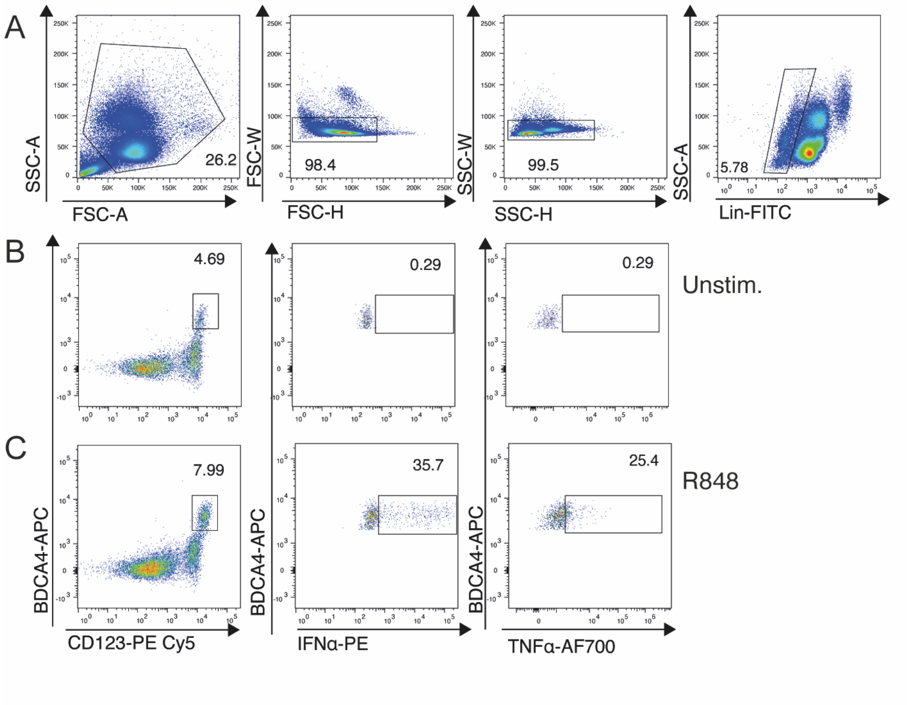


**Figure EV1. Gating strategy for flow cytometric analysis of cytokine-producing pDCs on freshly drawn PBMCs.**

A Gating strategy for analysis of human pDCs. pDCs were identified as Singlet, $CD14^{neg}CD19^{neg}CD123^{+}$ BDCA-$4^{+}$ cells.

B, C Freshly isolated PBMCs were stimulated for 5 hours with the TLR7/8 ligand R-848 (1.5 µg/ml) (C) or left untreatred (B), in the presence of brefeldin A for the final 3 hours, then harvested, surface stained, fixed, permeabilized, and intracellularly stained for IFN-$\alpha$ and TNF-$\alpha$.

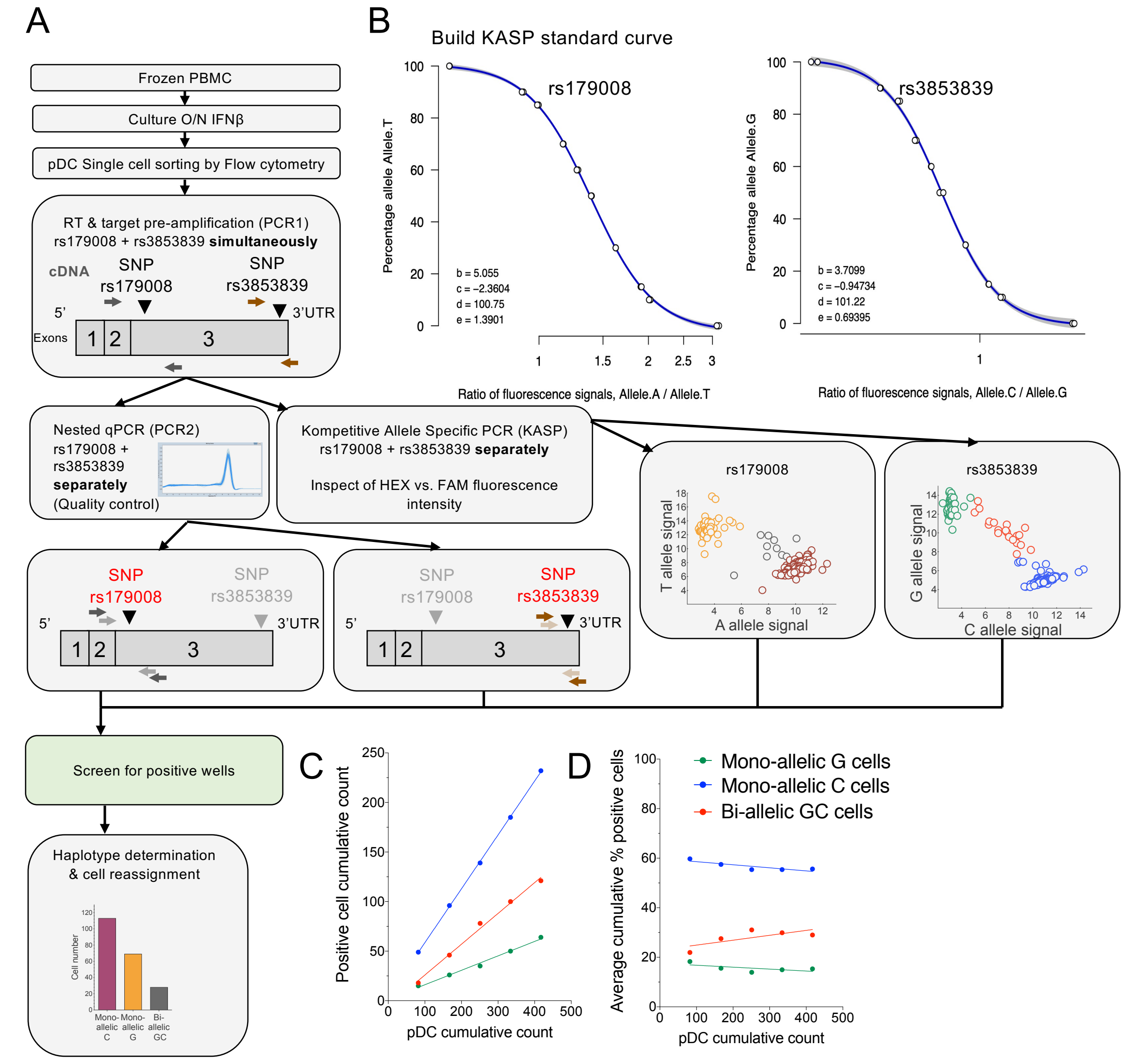


**Figure EV2 : Experimental flow chart for the single-cell RT-PCR-KASP analysis of *TLR7* allelic expression in pDCs from female donors heterozygous for the rs179008 (A/T) and/or rs3853839 (G/C) SNPs.**

A Workflow of the allele-of-origin analysis using two genetic markers observable in mature *TLR7* transcripts. Frozen PBMC were cultivated overnight with 1 ng/ml IFNβ. pDC were stained and single cell sorted by flow cytometry.

B Standard curve of the KASP assay for both SNPs.

C, D Absolut number or average frequency of monoallelic G (green) - C (blue) or biallelic (red) pDCs cumulative count among total pDCs from a UC female donor obtained by single-cell RT-PCR KASP.

Data information: (C,D) Cumulative results of sc-RT-PCR-KASP assays are shown from 2 experiments with 3 or 2 96-w plates performed with an average of 84 cells/plate analyzed in each experiment. Cumulative results of 5 single-cell sorted plates are shown.

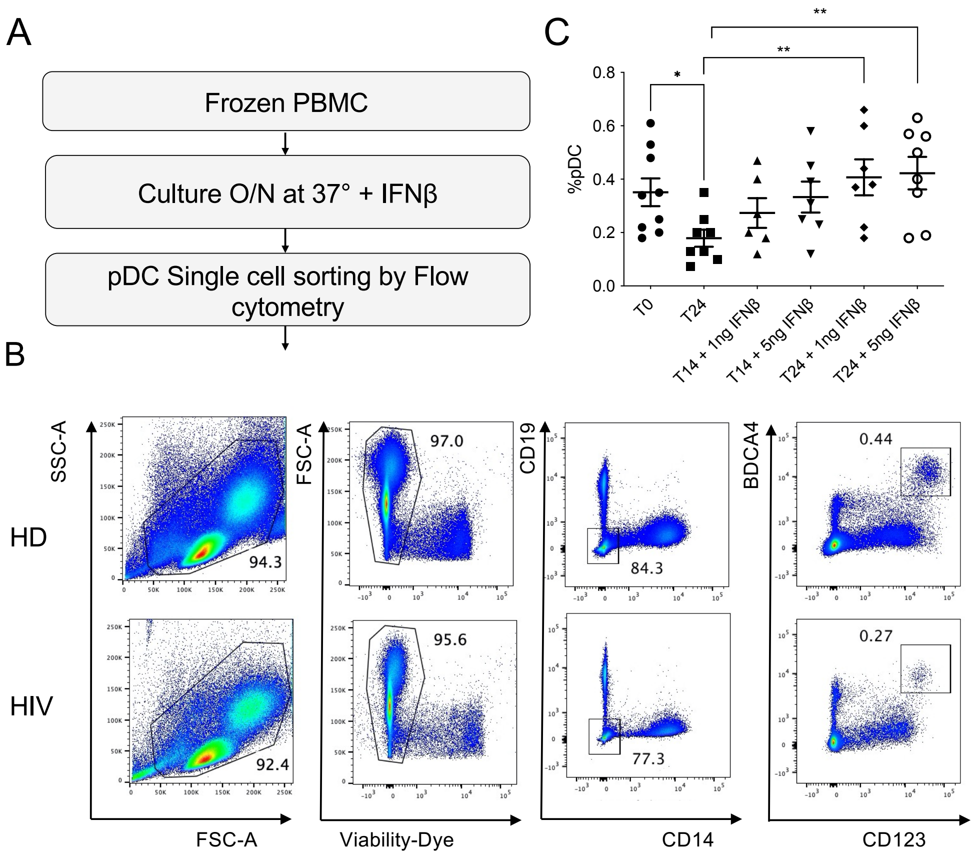


**Figure EV3. Gating strategy for single-cell sorting of pDCs from frozen PBMCs from HIV-1/ART or uninfected control female donors.**

A Frozen PBMCs were set in culture in complete medium in the presence of IFN-ß ON at 37°C, before labelling and cell-sorting in 96-well plate containing 4 µl of lysis-buffer.

B Gating strategy used to sort pDCs. pDCs were identified as Singlet, Live, $CD14^{neg}CD19^{neg}CD123^{+}$ BDCA-4$^{+}$ cells.

C In order to synchronize *TLR7* expression in pDCs and to optimize their survival in culture, IFN-ß was added in the culture medium. In panel C, the frequency of pDCs is shown in PBMCs from various donors at different time points of culture at 37°C in the presence of the indicated concentrations of IFN-ß.

Data information: (C) Error bars represent the mean ± SEM. statistical analysis was performed using a paired Student's *t*-test. *$P$ <0.05, **$P$<0.01.

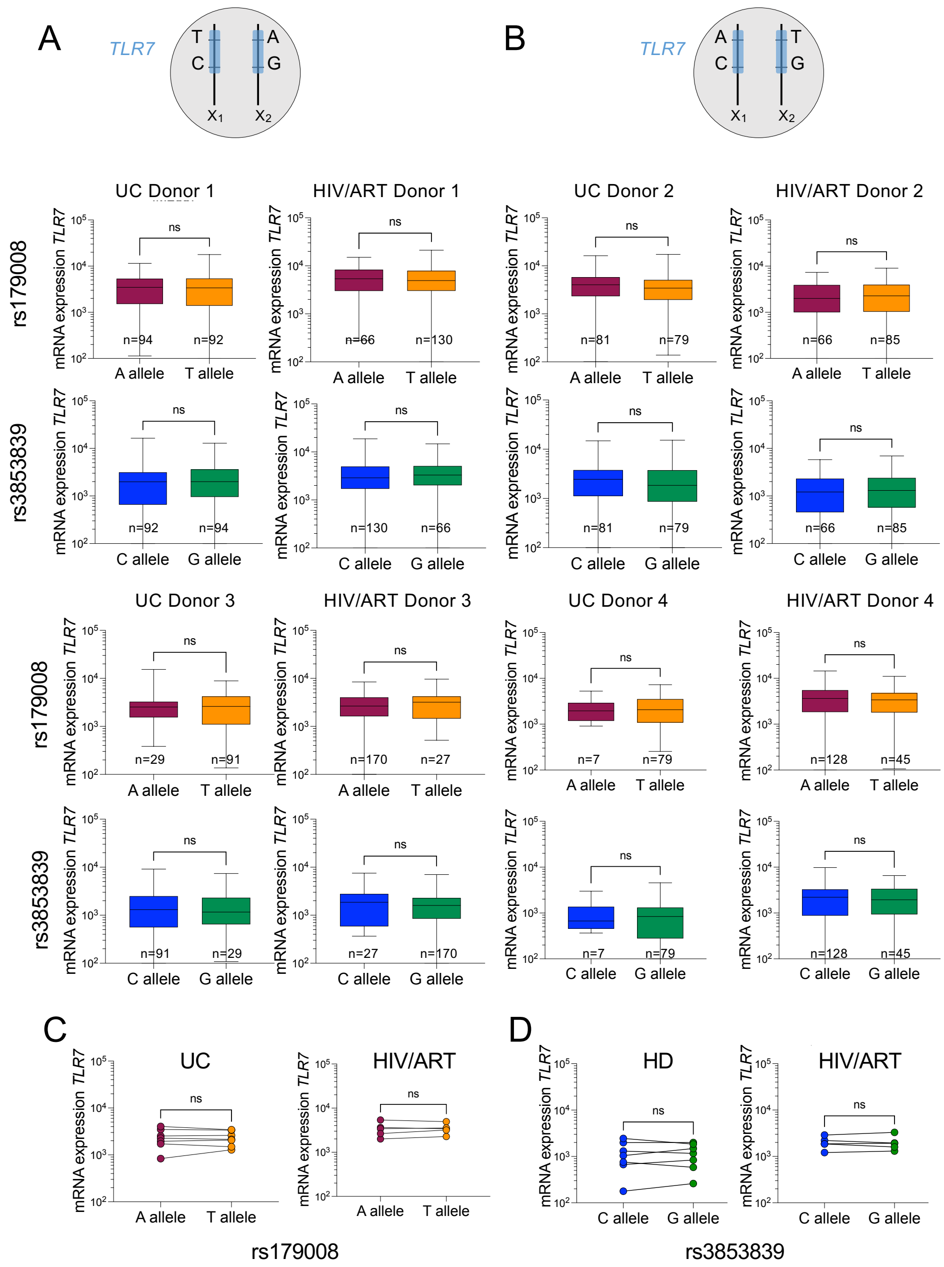

**Figure EV4: Mono-allelic expression of *TLR7* mRNA is not affected by the parental origin of the X chromosome nor by rs179008 or rs3853839 SNPs.**

A,B *TLR7* mRNA expression at single-cell resolution in monoallelic pDCs from representative uninfected controls (UC) and HIV/ART women heterozygous for both the SNPs rs179008 (A/T) and rs3853839 (C/G) whom carries the indicated haplotypes T-C/A-G (A) or A-C/T-G (B)

C, D To investigate the impact of each SNP on *TLR7* mRNA expression, the median values of *TLR7* mRNA expression at single-cell resolution for each allele were plotted from all UC donors (n=7) and HIV-1$^{+}$ women (n=5) as indicated, without distinction of haplotypes.

Data information: (A, B) Statistical differences in *TLR7* mRNA levels were assessed using the Mann and Whitney test. ns, not significant *P* >0.05. (C,D) Statistical differences in *TLR7* mRNA levels according to the rs179008 and rs385839 genotypes were analysed using the Wilcoxon signed rank test. ns, not significant *P* >0.05.
